# Sustained Cellular Immune Dysregulation in Individuals Recovering from SARS-CoV-2 Infection

**DOI:** 10.1101/2020.07.30.20165175

**Authors:** Jacob K. Files, Sushma Boppana, Mildred D. Perez, Sanghita Sarkar, Kelsey E. Lowman, Kai Qin, Sarah Sterrett, Eric Carlin, Anju Bansal, Steffanie Sabbaj, Dustin M. Long, Olaf Kutsch, James Kobie, Paul A. Goepfert, Nathan Erdmann

**Author notes:** Corresponding Authors: Nathan Erdmann, Bevill Biomedical Research Building Rm 512, 845 19^th^ Street S, Birmingham, AL 35294, USA, Paul Goepfert, Bevill Biomedical Research Building Rm 563, 845 19^th^ Street S, Birmingham, AL 35294, USA.

## Abstract

SARS-CoV-2 causes a wide spectrum of clinical manifestations and significant mortality. Studies investigating underlying immune characteristics are needed to understand disease pathogenesis and inform vaccine design. In this study, we examined immune cell subsets in hospitalized and non-hospitalized individuals. In hospitalized patients, many adaptive and innate immune cells were decreased in frequency compared to healthy and convalescent individuals, with the exception of B lymphocytes which increased. Our findings show increased frequencies of T-cell activation markers (CD69, Ox40, HLA-DR and CD154) in hospitalized patients, with other T-cell activation/exhaustion markers (CD25, PD-L1 and TIGIT) remaining elevated in hospitalized and non-hospitalized individuals. B cells had a similar pattern of activation/exhaustion, with increased frequency of CD69 and CD95 during hospitalization, followed by an increase in PD1 frequencies in non-hospitalized individuals. Interestingly, many of these changes were found to increase over time in non-hospitalized longitudinal samples, suggesting a prolonged period of immune dysregulation following SARS-CoV-2 infection. Changes in T-cell activation/exhaustion in non-hospitalized patients were found to positively correlate with age. Severely infected individuals had increased expression of activation and exhaustion markers. These data suggest a prolonged period of immune dysregulation following SARS-CoV-2 infection highlighting the need for additional studies investigating immune dysregulation in convalescent individuals.

## INTRODUCTION

Since the first reports in December 2019, Coronavirus Disease 2019 (COVID-19), caused by the novel virus, Severe Acute Respiratory Syndrome Coronavirus 2 (SARS-CoV-2), has spread worldwide and caused enormous public health and economic impacts (1, 2). Many patients remain asymptomatic or have mild symptoms (3), while others, particularly those with comorbidities, develop severe clinical diseases with atypical pneumonia and multiple system organ failure (4-6). To date, there have been over 16 million reported cases with 645,000 COVID-19 related deaths (7), and with no imminent vaccine, understanding of the immune pathology associated with patients of various clinical outcomes is urgently needed.

Several early studies have shown that acutely infected individuals develop lymphopenia (4, 8-11) and exhibit elevated expression of T-cell activation markers (12-14). Additionally, several groups have shown differential expression of T-cell exhaustion markers between severe and mild clinical cases (12, 13, 15). Studies have reported increased NKG2A levels in both CD8+ T cells and NK cells that return to baseline in convalescence (13), and increased monocyte frequencies in comparison to lymphocytes when using single-cell RNA sequencing analysis (16); however, these studies were limited in scope. Recent reports have described the presence of SARS-CoV-2-specific T cells in convalescent patients (17, 18), emphasizing the value of studying immune subsets in convalescent individuals who have recently overcome infection in order to better inform vaccine and therapeutic efforts.

Previous studies on Severe Acute Respiratory Syndrome Coronavirus (SARS) and Middle East Respiratory Syndrome Coronavirus (MERS) demonstrated increased immune system dysregulation with lymphopenia and inflammatory cytokine storm (19, 20). In patients infected by SARS, various studies investigated the T cell and B cell subsets(21-23). Early investigations in SARS-CoV-2 (24, 25) and more recent large cohort studies have described immune perturbations in acutely infected hospitalized individuals with severe infection (26, 27). In acute viral infections such as influenza, several groups have shown evidence of prolonged immune activation during the convalescent phase that correlated with worse clinical outcomes (28-30). Studies investigating antibody responses in MERS were found to be undetectable in patients who had mild illness following infection (31). These findings highlight the need for studies investigating the immune system of both hospitalized and non-hospitalized SARS-CoV-2 individuals. Here, we evaluate immune cell subsets of SARS-CoV-2 infected individuals and identified dysregulated immune cells in both hospitalized and non-hospitalized individuals over time.

## RESULTS

### Differential Immune Cell Subset Frequencies in Hospitalized and Non-Hospitalized Individuals

Others previously described that SARS-CoV-2 infected individuals, especially those with severe infection, have pronounced lymphopenia compared to healthy and convalescent individuals (4, 8, 9). However, most of these studies were done in hospitalized patients and only evaluated a few cell types. Here, we obtained samples and clinical data from a cohort of hospitalized COVID-19 patients (“Hospitalized”, n=46) and a cohort of non-hospitalized individuals who had recovered from confirmed COVID-19 infection (“Non-hospitalized”, n=39). These groups were compared to healthy, COVID-19 negative controls (“Healthy”, n=20). An overview of the cohort demographics is shown in **Table 1**. Importantly, most individuals in the hospitalized group (n=36) were viremic and hospitalized at the time of sample collection; however, a minority (10 of 46) were asymptomatic for at least 3 consecutive days and were at least 7 days past initial diagnosis at the time of initial sample collection and therefore could be classified as convalescent, but were still included in the hospitalized group due to disease severity. All individuals in the non-hospitalized group were convalescent at the time of sample collection. As shown in **Table 1**, there were differences between age, race, and comorbidities. Many of these differences reflect the nature of the COVID-19 pandemic, with more severe infection being associated with older age, African-American race, and those with pre-existing comorbidities.

**Table 1:**
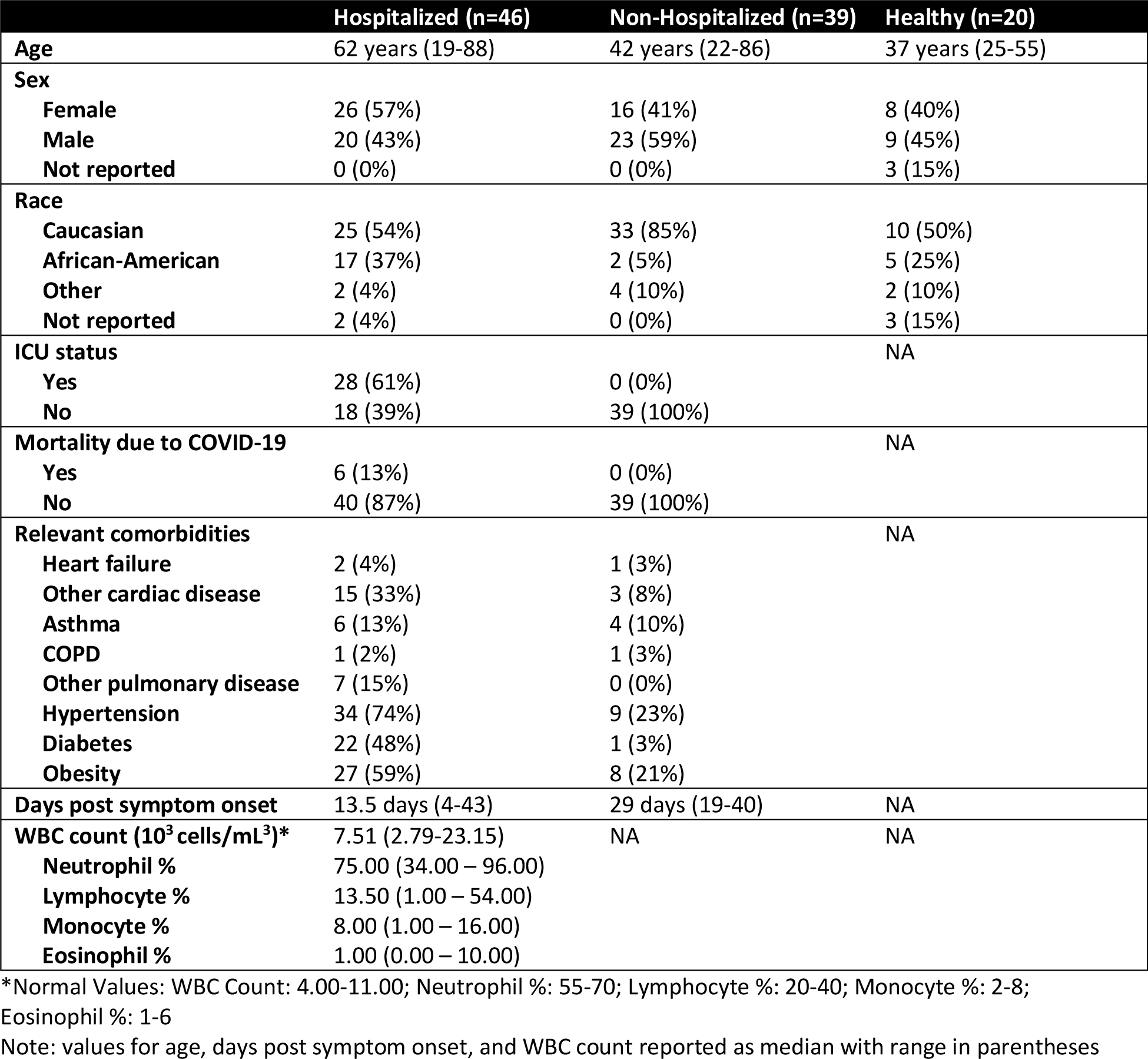
Cohort Demographics and Clinical Information

As previously described (32), we found that hospitalized individuals in our study displayed normal WBC counts but distinctly lower numbers of lymphocytes (**Table 1**). To further assess immune cells that remain during SARS-CoV-2 infection, we utilized a general immunophenotyping flow cytometry panel to evaluate the proportion of specific immune cell subsets within the total CD45+ cell population in each individual (overview shown in **Figure 1A**). While there was a decrease in the lymphocyte counts, we observed no significant decrease in proportion of CD4+ T cells in the total CD45+ population (**Figure 1B**). There was an increase in CD8+ T cell frequencies in the non-hospitalized group compared to the hospitalized group (**Figure 1C**, p=0.003). In contrast, B-cell frequencies were decreased in non-hospitalized individuals in comparison to our healthy and hospitalized groups (**Figure 1D**, p=0.002 and p<0.001, respectively).

**Figure 1:**
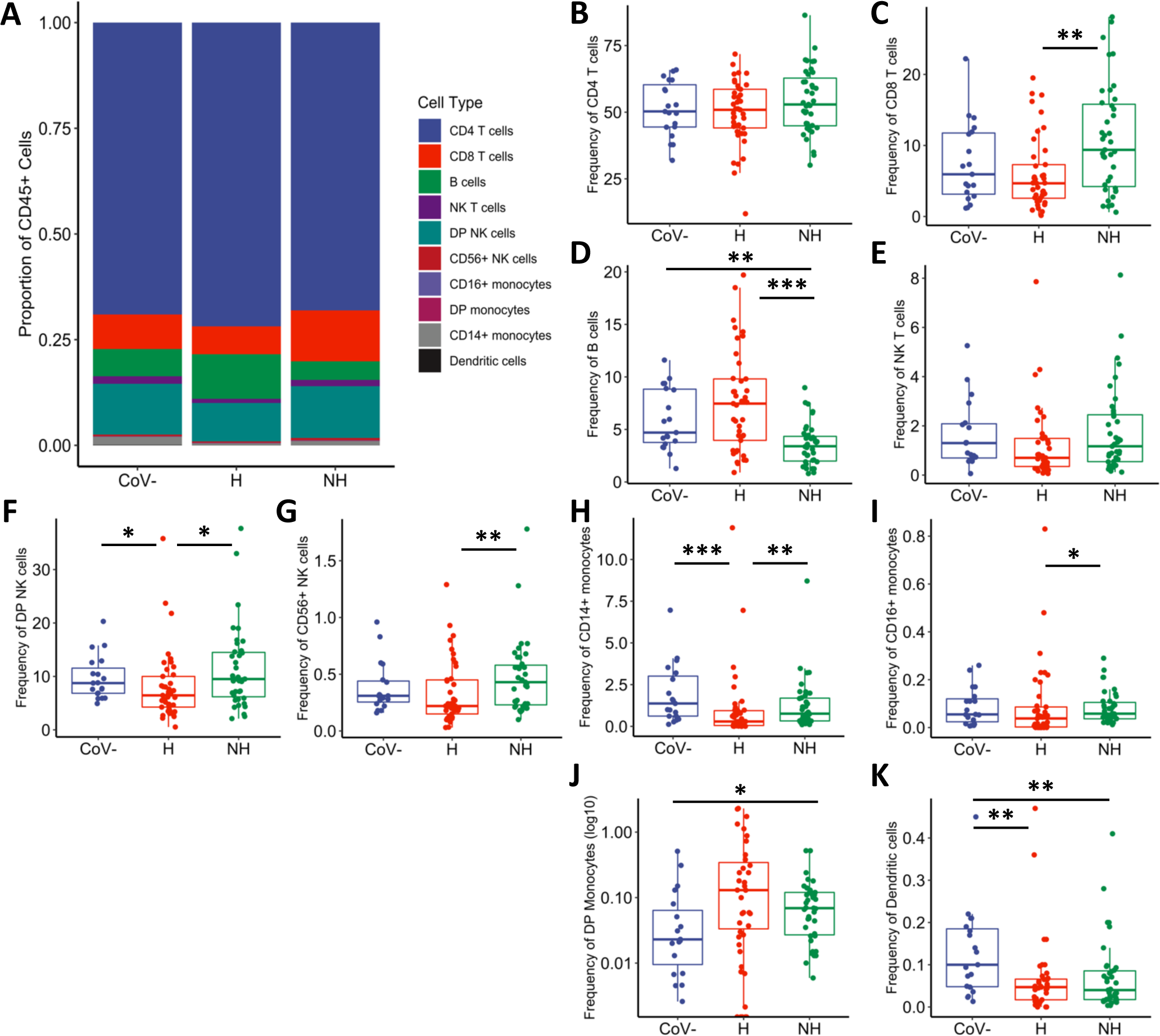
Differential frequencies of immune cell subsets in hospitalized and non-hospitalized individuals. Staining of isolated PBMCs from healthy (CoV-, n=19), hospitalized (H, n=41), and non-hospitalized (NH, n=39) samples, showing immune cell subsets as a frequency of the total CD45+ population. **A)** Overview of all immune cell subsets, with a more in-depth look at **B)** CD4+ and **C)** CD8+ T cells; **D)** B cells; **E)** NK T cells; **F)** CD56+CD16+ and **G)** CD56+CD16-NK cells; **H)** CD16+, **I)** CD14+CD16+, and **J)** CD14+ monocytes; and **K)** dendritic cells. Boxplots indicate median, IQR, and 95% confidence interval. P-values determined by Wilcoxon ranked sum tests and are indicated as follows: *p≤0.05, **p≤0.01, ***p≤0.001.

We investigated the innate immune cell subsets including NK T cells, NK cells, monocytes, and dendritic cells, which have been shown to play a protective role during other acute viral infections, including influenza A (33). Interestingly, many of these subsets showed decreased frequencies in hospitalized patients and returned to baseline in non-hospitalized individuals. There was a trend towards decreased NK T cells in the hospitalized group compared to healthy and non-hospitalized (**Figure 1E**, p=0.057 and p=0.057, respectively). Similarly, hospitalized individuals had decreased double positive (DP) CD56+CD16+ NK cells (**Figure 1F**, compared to healthy: p=0.023 and compared to non-hospitalized: p=0.011) and CD56+CD16-NK cells (**Figure 1G**, compared to non-hospitalized: p=0.009). There were decreased frequencies of CD14+ monocytes in hospitalized individuals when compared to healthy and non-hospitalized (**Figure 1H**, p<0.001 and p=0.004, respectively), with decreased frequencies in the CD16+ monocyte population of hospitalized over non-hospitalized as well (**Figure 1I**, p=0.013). In contrast, the amount of double positive CD16+CD14+ monocytes increased in frequency in the non-hospitalized group above healthy controls (**Figure 1J**, p=0.033). Dendritic cell frequency decreased in both our hospitalized and non-hospitalized samples were decreased as compared to healthy (**Figure 1K**, p=0.003 and p=0.007 respectively). These data show altered immune cell frequencies in hospitalized individuals, and while some of these observed perturbances are missing in the non-hospitalized individuals, decreased frequencies of DP monocytes and dendritic cells suggest a degree of sustained immune dysfunction in both groups of SARS-CoV-2-infected individuals.

### Immune Dysregulation in Hospitalized and Non-hospitalized Samples

Prior findings suggest that T cells upregulate activation and exhaustion markers during acute COVID-19 infection, especially in severe infection (12-15, 26, 27). Here, we examine T-cell phenotypes in infection and convalescence, separating our cohort based on severity of infection into individuals who were hospitalized and non-hospitalized. We measured surface-level expression of the activation markers CD69, Ox40, CD38, CD137, and CD154. We also analyzed the T-cell exhaustion markers TIGIT, PDL1, PD1 and Tim3. Notably, PD1 and Tim3, while often classified as exhaustion markers, have also been shown to be activated during other acute infections (34, 35). Finally, we stained for CD27 and CD28 as loss of these markers has been found to represent a loss of differentiated memory T cells, suggesting a more senescent phenotype (36).

An overview of the relative frequencies of all cell markers within CD4+ T cells are shown in the heatmap in **Supplemental Figure 1A**. In hospitalized individuals, we observed elevated frequencies over healthy and non-hospitalized groups with respect to CD69 (**Figure 2A**, p<0.001 and p<0.001, respectively), Ox40 (**Figure 2B**, p<0.001 and p<0.001, respectively) and PD1 (**Figure 2C**, p<0.001 and p=0.005, respectively). The markers HLA-DR, CD154, and Tim3 were elevated in hospitalized over non-hospitalized individuals (**Figures 2D-2F**, p=0.009, p=0.028 and p=0.047, respectively). Collectively, these observations identify markers elevated during severe infection, but then return to baseline with resolution of symptoms or remain normal in mild infection. Further, two exhaustion markers were elevated in both hospitalized and non-hospitalized samples when compared to healthy: TIGIT (**Figure 2G**, p<0.001 and p<0.001, respectively) and PDL1 (**Figure 2H**, p= 0.007 and p=0.003, respectively). Meanwhile, the frequency of CD4+ T cells expressing CD38 were significantly lower in hospitalized patients in comparison to non-hospitalized (**Figure 2I**, p=0.031). In summary, expression of some markers (CD69, Ox40, PD1, HLA-DR, CD154 and Tim3) on CD4+ T cells appear elevated in hospitalized SARS-CoV-2 infected individuals, while they appear to be similar to baseline in non-hospitalized individuals. Other activation and exhaustion markers such as TIGIT and PDL1 remain elevated during the convalescent stage post infection.

**Figure 2:**
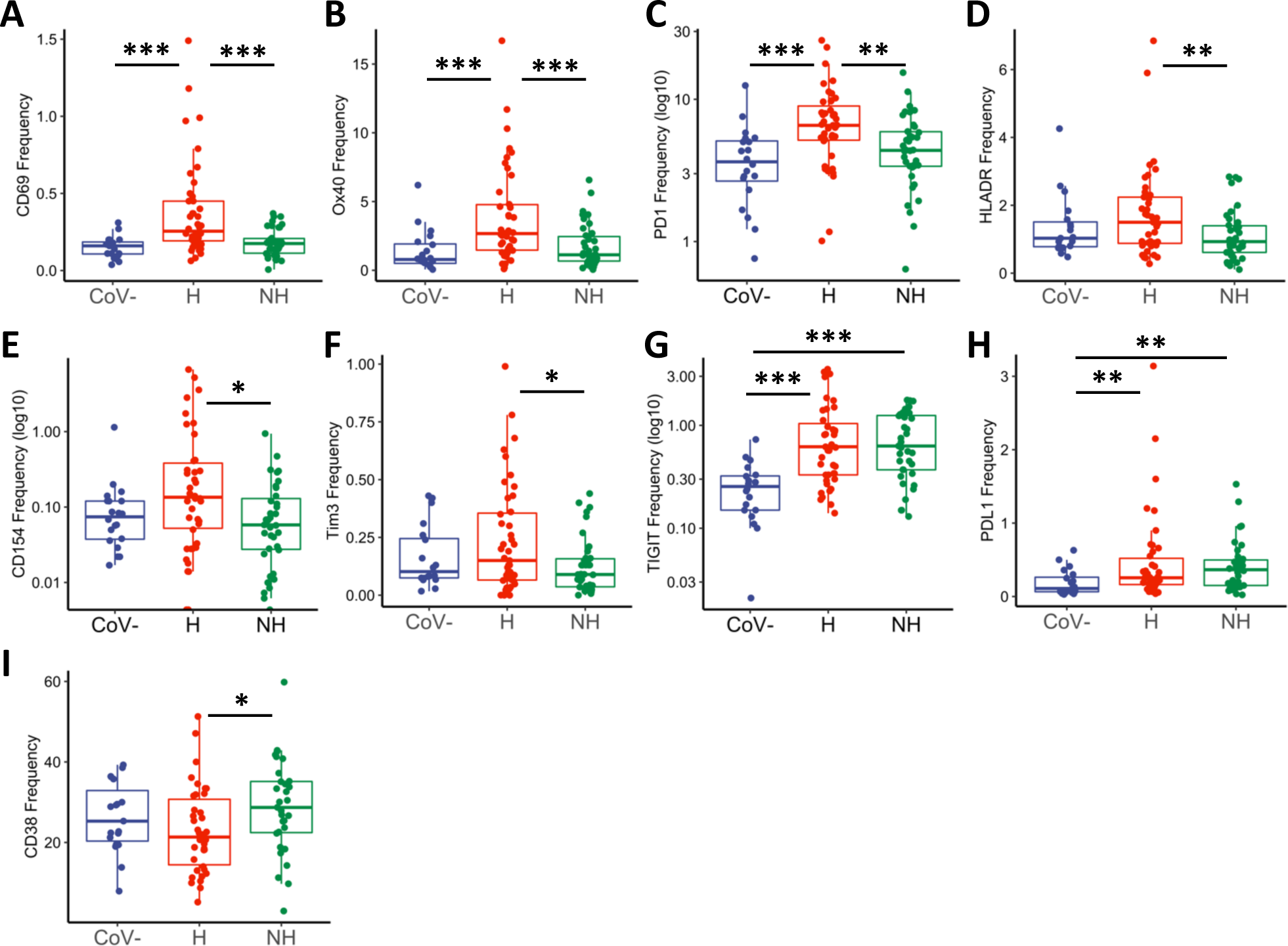
CD4+ T cell activation and exhaustion in hospitalized and non-hospitalized individuals. Frequency of CD4+ T cells expressing a given activation or exhaustion marker. **A-F**) Markers that are elevated in hospitalized patients and similar to baseline in non-hospitalized. **G-I)** Markers that remain elevated in both hospitalized and non-hospitalized. Boxplots indicate median, IQR, and 95% confidence interval. P-values determined by Wilcoxon rank sum tests and are indicated as follows: *p≤0.05, **p≤0.01, ***p≤0.001. Healthy: CoV-(n_CoV-_= 20), Hospitalized: H (n_H_= 46), Non-hospitalized: NH (n_NH_= 39).

We next investigated the frequencies of T-cell activation and exhaustion markers in CD8+ T cells (relative frequencies of all markers shown in **Supplemental Figure 1B**). We found that the activation markers CD69 and CD38 were significantly elevated in hospitalized individuals in comparison to healthy (**Figures 3A-3B**, p<0.001 and p=0.002, respectively) and non-hospitalized groups (**Figure 3A-3B**, p<0.001 and p<0.001, respectively). In addition, frequencies of Ox40, CD154, and HLA-DR were all higher in hospitalized individuals than non-hospitalized (**Figures 3C-3E**, p=0.033, p=0.013, and p=0.005, respectively). The exhaustion marker Tim3 was higher in the hospitalized than the non-hospitalized group (**Figure 3F**, p=0.004), and PDL1 was higher in hospitalized over non-hospitalized and healthy groups (**Figure 3G**, p=0.001 and p<0.001,respectively). Overall, expression of these activation and exhaustion markers indicates more severe immune dysregulation of CD8+ T cells in the hospitalized group.

**Figure 3:**
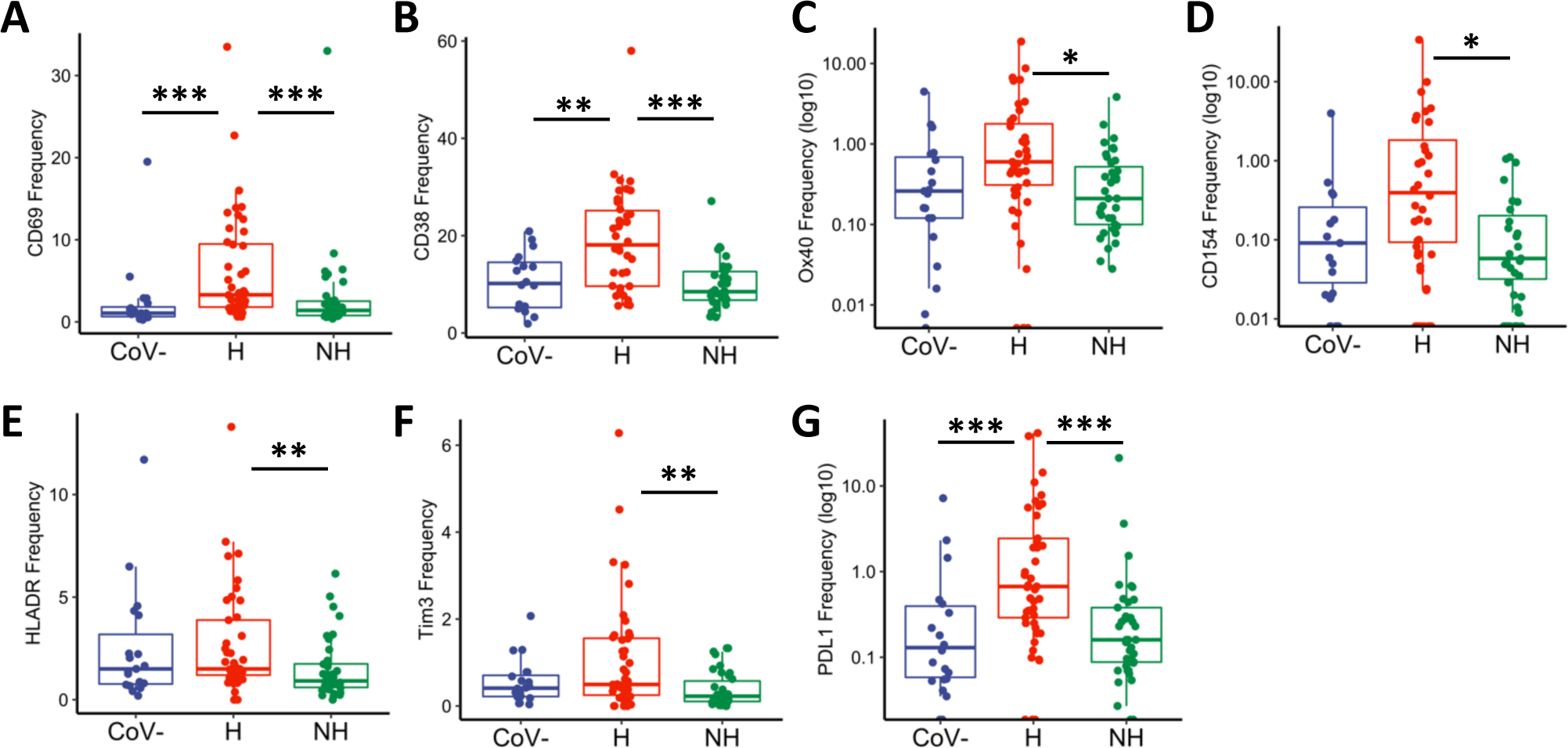
CD8+ T cell activation and exhaustion in hospitalized and non-hospitalized individuals. Frequency of CD8+ T cells expressing a given activation or exhaustion marker. **A-G**) Markers that are elevated in hospitalized over non-hospitalized. Boxplots indicate median, IQR, and 95% confidence interval. P-values determined by Wilcoxon rank sum tests and are indicated as follows: *p≤0.05, **p≤0.01, ***p≤0.001. Healthy: CoV-(n_CoV-_= 20), Hospitalized: H (n_H_= 46), Non-hospitalized: NH (n_NH_= 39).

To help characterize the B lymphocyte population in this cohort, we looked at the activation markers CD69, CD95, and HLA-DR. Fc receptor-like 4 (FcRL4) was also measured, as it is upregulated on B cells of lymphoid tissue, increased in the periphery during some chronic infections and autoimmune diseases, and is associated with an exhausted B-cell phenotype (37-42). We also examined the frequencies of the exhaustion marker PD1, and the memory marker, CD27. A summary of B-cell marker expression is shown in **Supplemental Figure 1C**. Similar to observations in our CD4+ and CD8+ T cell subsets, we found an increased frequency of the activation markers CD69 and CD95 in hospitalized individuals over non-hospitalized (**Figures 4A and 4B**, p=0.039 and p=0.034, respectively), while CD69 was also elevated in both hospitalized and non-hospitalized groups compared to healthy controls (**Figure 4A**, p<0.001 and p<0.001, respectively). B-cell expression of CD95 was also increased in hospitalized individuals over healthy controls (**Figure 4B**, p=0.021). B-cell expression of HLA-DR trended toward decrease in hospitalized individuals, while CD27 is significantly increased in non-hospitalized individuals over healthy controls (**Figures 4C and 4D**, p=0.051 and p=0.004, respectively). Finally, the exhaustion marker PD1 is elevated in non-hospitalized individuals over hospitalized patients and healthy controls (**Figure 4E**, p=0.026 and p=0.016, respectively). These data indicate that B cells are dysregulated in both hospitalized and non-hospitalized COVID-19 patients, similar to what we have described for T cells.

**Figure 4:**
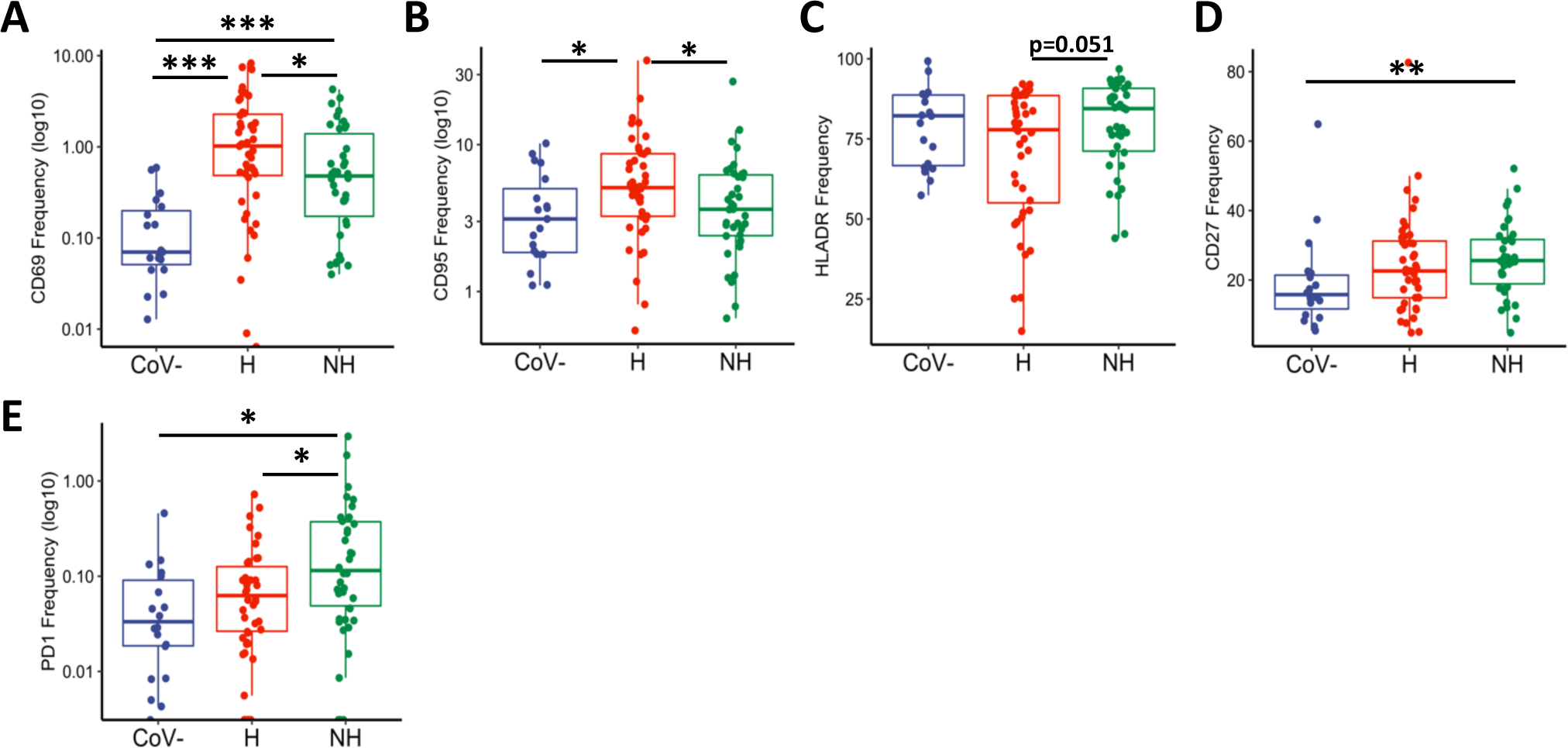
B cell activation and exhaustion in hospitalized and non-hospitalized individuals. Frequency of B cells expressing a given activation or exhaustion marker. **A-B**) CD95 and CD69 frequencies are elevated in hospitalized, while **C-D**) HLA-DR and CD27 frequencies are elevated in non-hospitalized samples. **E**) PD1 frequencies remain elevated in non-hospitalized. Boxplots indicate median, IQR, and 95% confidence interval. P-values determined by Wilcoxon rank sum tests and are indicated as follows: *p≤0.05, **p≤0.01, ***p≤0.001. Healthy: CoV-(n_CoV-_= 20), Hospitalized: H (n_H_= 46), Non-hospitalized: NH (n_NH_= 39).

### Longitudinal analysis shows sustained immune dysregulation in non-hospitalized convalescent individuals

We next performed longitudinal analyses comparing the expression of activation and exhaustion markers over time. We did this in two ways: 1) observing changes in marker frequencies over time, defined as days post-symptom onset (median = 29 days; range 19-40), utilizing all non-hospitalized samples with a recorded date post-symptom onset (n=23); and 2) directly comparing the frequencies of these markers between Visit 1 and Visit 2 for those non-hospitalized patients with samples collected at two sequential timepoints (n=25).

Interestingly, when investigating changes in activation and exhaustion markers we observed that CD4+ T-cell expression of HLA-DR increased over time in our non-hospitalized samples (**Figure 5A**, p=0.022 by days post symptom onset and p=0.013 by visit 1/visit 2). Similarly, Ox40 and Tim3 frequencies increased over time (**Figures 5B and 5C**), with trending relationships based on days post symptom onset analyses (p=0.056 and p=0.055, respectively) and significant relationships based on visit 1/visit 2 analyses (p=0.001 and p=0.031, respectively). We also observed an increase in CD69 expression between visit 1 and visit 2 (**Supplemental Figure 2A**, p=0.006), and an increase in CD137 based on days post symptom onset (**Supplemental Figure 2B**, p=0.027), though these markers were not significantly upregulated over both methods of analysis. In contrast, frequencies of PDL1 decreased by days post symptom onset and between visit 1 and visit 2 (**Figure 5D**, p=0.028 and p=0.016, respectively). Taken together, these data show evidence that CD4+ T cells in non-hospitalized patients continue to express, and even upregulate, T-cell activation and exhaustion markers.

**Figure 5:**
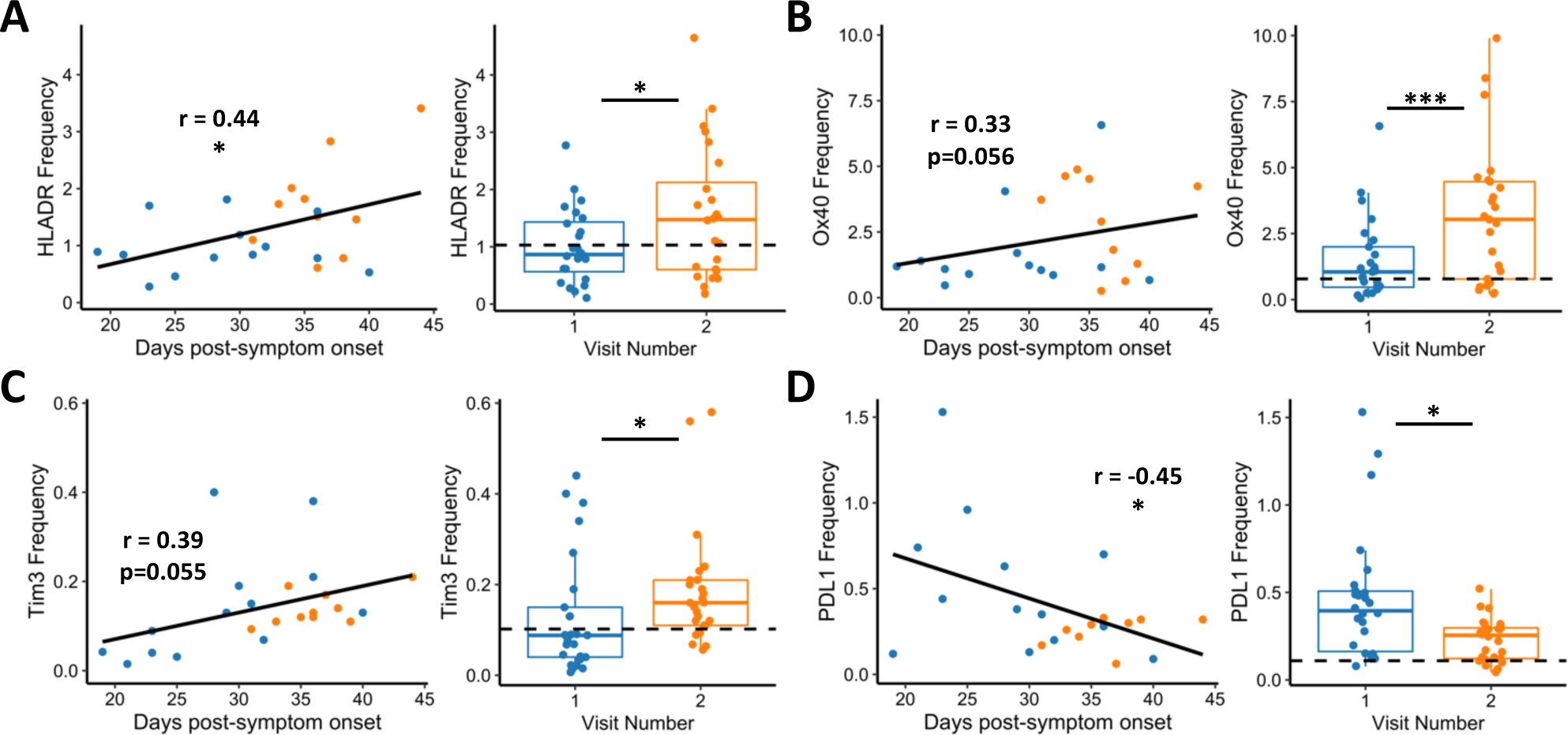
CD4+ T cell activation and exhaustion over time in non-hospitalized individuals. **A-C**) HLA-DR, Ox40, and Tim3 frequencies increased over time, while **D)** PDL1 frequency decreased over time in non-hospitalized patients. Plots on **left** show days post-symptom onset versus frequency (n=23, p-values determined by mixed effect model and relationship represented by linear regression; Blue: visit 1, Orange: visit 2). Plots on **right** show paired analysis of first versus second visits (n=25, p-values determined by paired Wilcoxon signed-rank test). Dotted line shows median values of Healthy samples as baseline. Boxplots indicate median, IQR, and 95% confidence interval. All p-values are indicated as follows, unless otherwise stated: *p≤0.05, **p≤0.01, ***p≤0.001.

When investigating CD8+ T-cell marker frequencies in non-hospitalized individuals longitudinally, we observed increased expression of the activation marker HLA-DR based on days post-symptom onset and visit 1/visit 2 analyses (**Figure 6A**; p=0.052 and p=0.031, respectively). Other activation markers also had an increased frequency at the later timepoint, including CD69 and CD154 (**Supplemental Figure 2C-D**, p<0.001 and p=0.007 respectively). Additionally, we found that CD8 T-cell expression of exhaustion markers increased in non-hospitalized individuals over time when looking at days post-symptom onset and visit 1/visit 2 analyses for TIGIT and PDL1 (**Figure 6B and 6C**, p=0.006, p=0.003, p=0.051 and p=0.009, respectively), as well as Tim3 (**Supplemental Figure 2E**, p=0.036). We also observed significantly decreased frequencies of CD27 and CD28 based on days post-symptom onset and visit 1/visit 2 analyses, albeit with low correlation coefficients (**Figure 6D and 6E**, p=0.039, p=0.014, p=0.035 and p=0.009, respectively). In summary, we show that the frequency of several activation and exhaustion markers in CD8+ T cells may initially be lower in non-hospitalized individuals in comparison to the hospitalized group; surprisingly, many of these markers appear to increase over time in the convalescent phase in these individuals.

**Figure 6:**
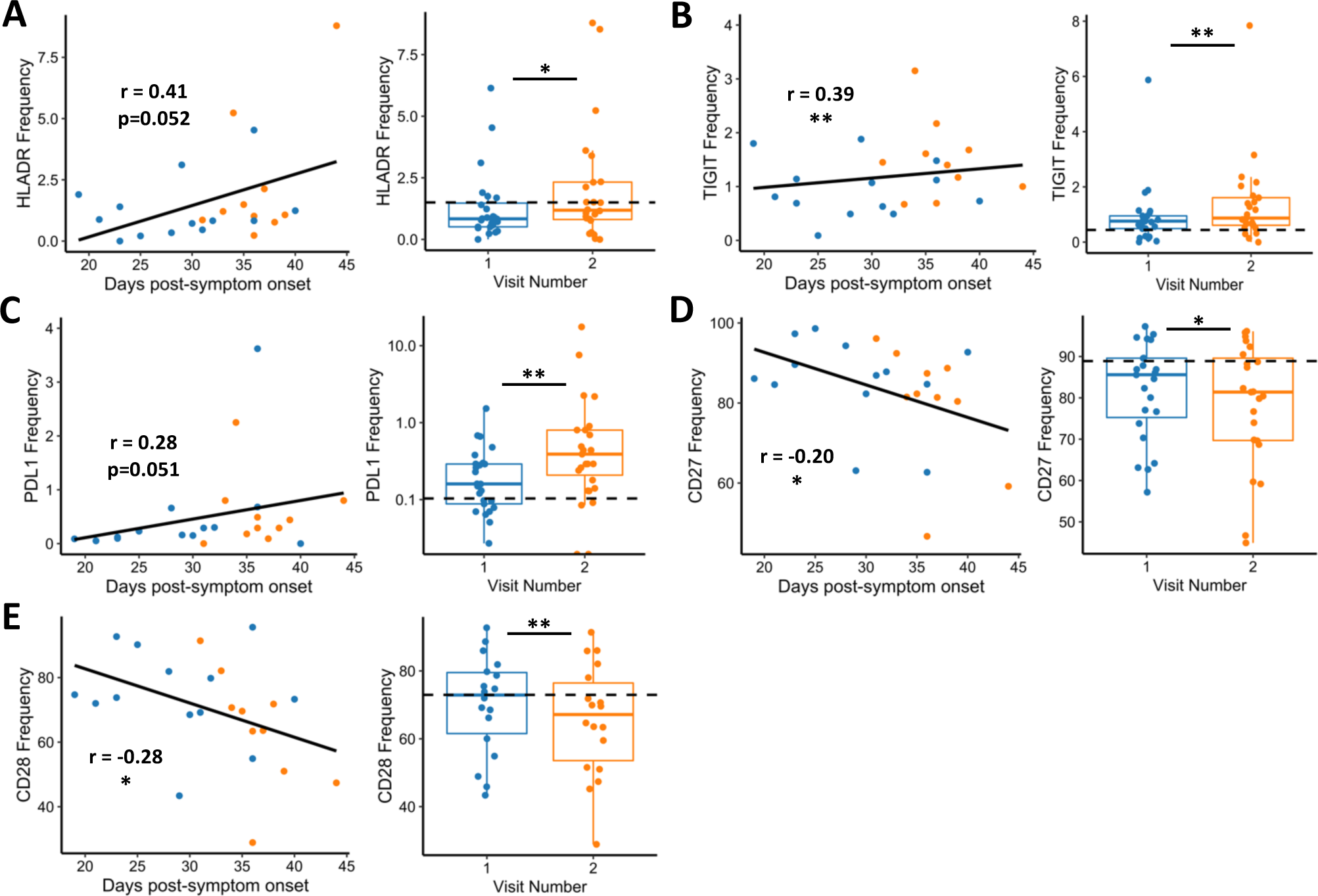
CD8+ T cell activation and exhaustion over time in non-hospitalized individuals. **A-C**) Expression of HLA-DR, TIGIT, and PDL1 increases over time in non-hospitalized individuals, while **D-E**) CD27 and CD28 frequencies decrease over time. Plots on **left** show days post-symptom onset versus frequency (n=23, p values determined by mixed effect model, relationship represented by linear regression; Blue: visit 1, Orange: visit 2). Plots on **right** show paired analysis of first versus second convalescent visits (n=25, p values determined by paired Wilcoxon signed-rank test). Dotted line shows median values of Healthy samples as baseline. Boxplots indicate median, IQR, and 95% confidence interval. All p-values are indicated as follows: *p≤0.05, **p≤0.01, ***p≤0.001.

Longitudinal investigation of non-hospitalized patients did not identify B-cell markers that significantly differed by both methods of analysis (days post symptom onset and visit 1 versus visit 2 comparison). However, B cells did express higher frequencies of FcRL4 and CD95 based on days post-symptom onset, and HLA-DR frequencies increased at the visit 2 timepoint (**Supplementary Figure 3A-C**, p=0.010, p=0.023, p=0.045, respectively). Meanwhile, CD27 frequency decreased over days post symptom onset, with decreased PD1 frequencies in the visit 2 group (**Supplementary Figure 3D and 3E**, p<0.001 and p=0.005, respectively). Overall, these data begin to suggest that the B cell population remains activated in non-hospitalized patients well into the convalescent period.

### Age Impacts T-Cell Activation and Exhaustion Markers in Hospitalized and Non-hospitalized Individuals

Since older SARS-CoV-2 patients have higher morbidity and mortality rates (43), we investigated differences related to age in each of our groups. By analyzing samples at the visit 1 timepoint in the hospitalized group, we found a positive correlation between age and CD8+ T-cell expression of CD69 (**Supplemental Figure 4A**, p=0.009), as well as a negative correlation between age and CD8 expression of CD27 and CD28 (**Supplemental Figure 4B and 4C**, p=0.010 and p=0.003, respectively). We observed a positive correlation between age and frequencies of CD69, CD95, and FcRL4 within the B-cell population of our hospitalized individuals (**Supplemental Figure 4D-4F**; p=0.024, p=0.024, and p=0.014, respectively). Analysis of CD4+ T cells and age in hospitalized individuals had no significant findings. We observed no differences between age and marker frequencies in healthy controls across all subsets.

We next investigated any correlations of age and frequency of activation and exhaustion markers in the non-hospitalized group. Our group and others have reported that elderly individuals form suboptimal immune responses following vaccination and infection (44). In this study, within the CD4+ T-cell compartment, we observed increased frequencies of PD1, TIGIT, and HLA-DR that correlated positively with age (**Figure 7A-C**, p=0.002, p=0.032 and p=0.022, respectively). We also observed a loss of CD28 expression in elderly individuals (**Figure 7D**, p=0.027). Similarly, in our CD8+ T-cells, there were increased PD1, HLA-DR, and TIGIT frequencies that correlated positively with age (**Figure 7E-7G**, p<0.001, p=0.007, and p=0.008, respectively), while expression of CD27 and CD28 had a negative correlation with age (**Figure 7H and 7I**, p<0.001 and p=0.010, respectively). Finally, there was a decrease in CD27+ B cells with age (data not shown, p=0.007). Of note, T cell and B cell expression of CD27 and CD28 has been shown to decline in the elderly by several groups (45). While more in-depth studies investigating T-cell function with age in SARS-CoV-2 infected individuals are necessary, these data found that T-cell immune dysregulation following SARS-CoV-2 infection is more pronounced in older non-hospitalized patients, suggesting that older individuals may have an impaired ability to form SARS-CoV-2-specific memory responses.

**Figure 7:**
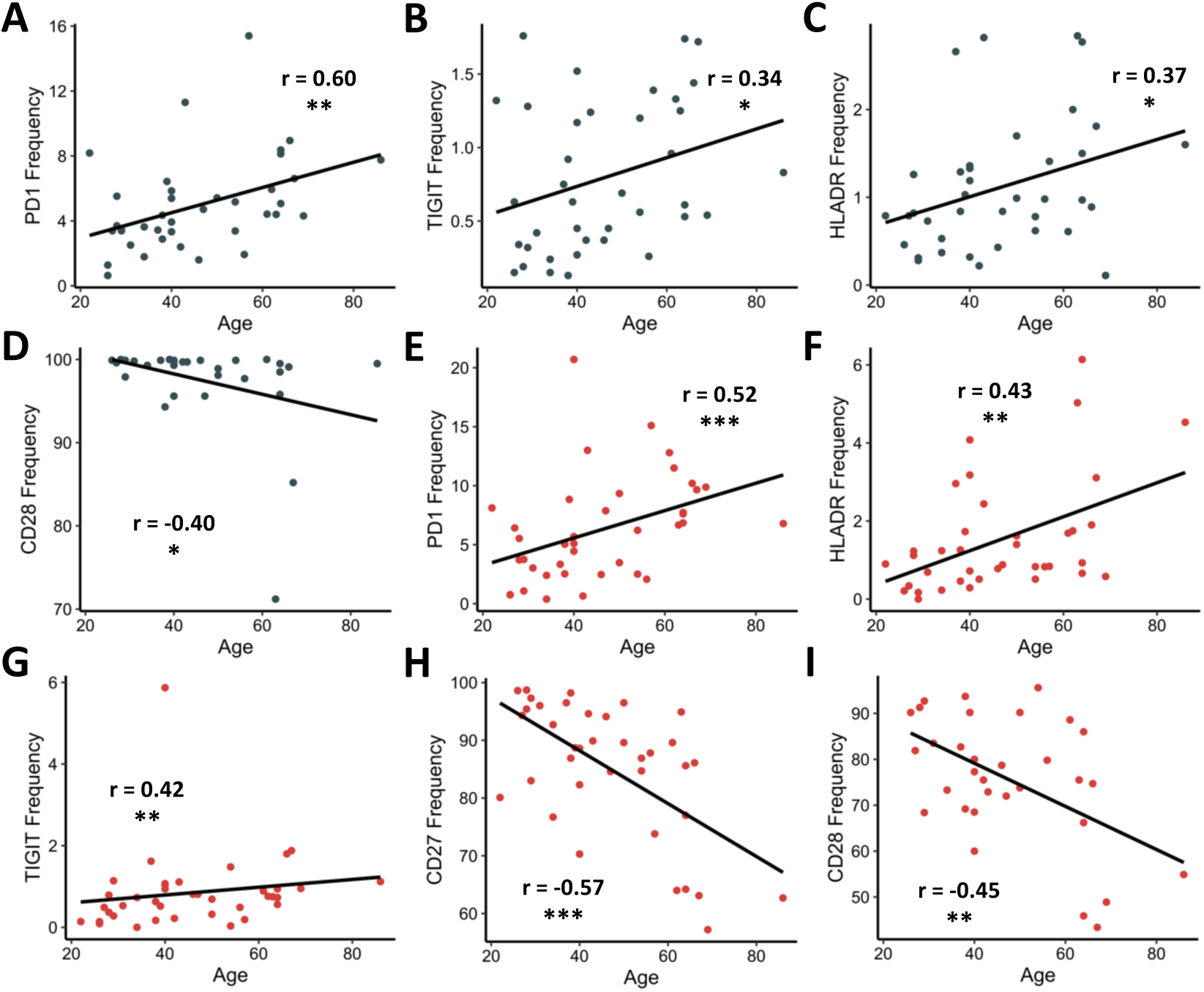
Correlations between T-cell marker frequencies and age in non-hospitalized individuals. **A-C**) PD1, TIGIT, and HLA-DR frequencies on CD4+ T cells (grey dots) increase with age. **D**) CD28 frequencies on CD4+ T cells decrease with age. **E-G**) PD1, HLA-DR, and TIGIT frequencies on CD8+ T (red dots) cells increase with age. **H-I**) CD27 and CD28 on CD8+ T cells decrease with age. R and p-values determined by Spearman rank correlation tests and are indicated as follows: *p≤0.05, **p≤0.01, ***p≤0.001; n = 39.

### Hospitalized ICU Patients have Increased T-cell and B-cell Dysregulation

Finally, we evaluated whether severity of illness impacted expression of activation and exhaustion markers within our hospitalized group as several other reports have found increased expression of activation/exhaustion markers in severe infection (12-15, 26, 27). We stratified samples from our hospitalized group into two groups: ICU (n=26) and non-ICU patients (n=10). (Note: 10 samples from the hospitalized group were collected in the convalescent period and thus were excluded in this analysis.) Our analysis found that ICU patients had increased CD69 expression on CD4+ T cells (**Figure 8A**, p=0.004), and CD38 frequencies were elevated in both CD4+ and CD8+ T cells (**Figures 8B and 8C**, p=0.049 and p=0.025, respectively), similar to recent results (12-15, 26, 27). In addition, the exhaustion marker PDL1 was found to be elevated in ICU patients (**Figure 8D**, p=0.018). Interestingly, the marker CD137 was found to be decreased in frequency in ICU patients (**Figure 8E**, p=0.002). Of note, CD137 has been found to play an important role in acute infection within mice (46). B cells from ICU patients had a trending increase in CD95 frequencies, significant increase in CD27 frequencies, while having decreased expression of HLA-DR (**Figures 8F-8H**, p=0.055, p=0.029, and p=0.045, respectively). In summary, these findings better define the dysfunctional immune response observed in severe SARS-CoV-2 infection.

**Figure 8:**
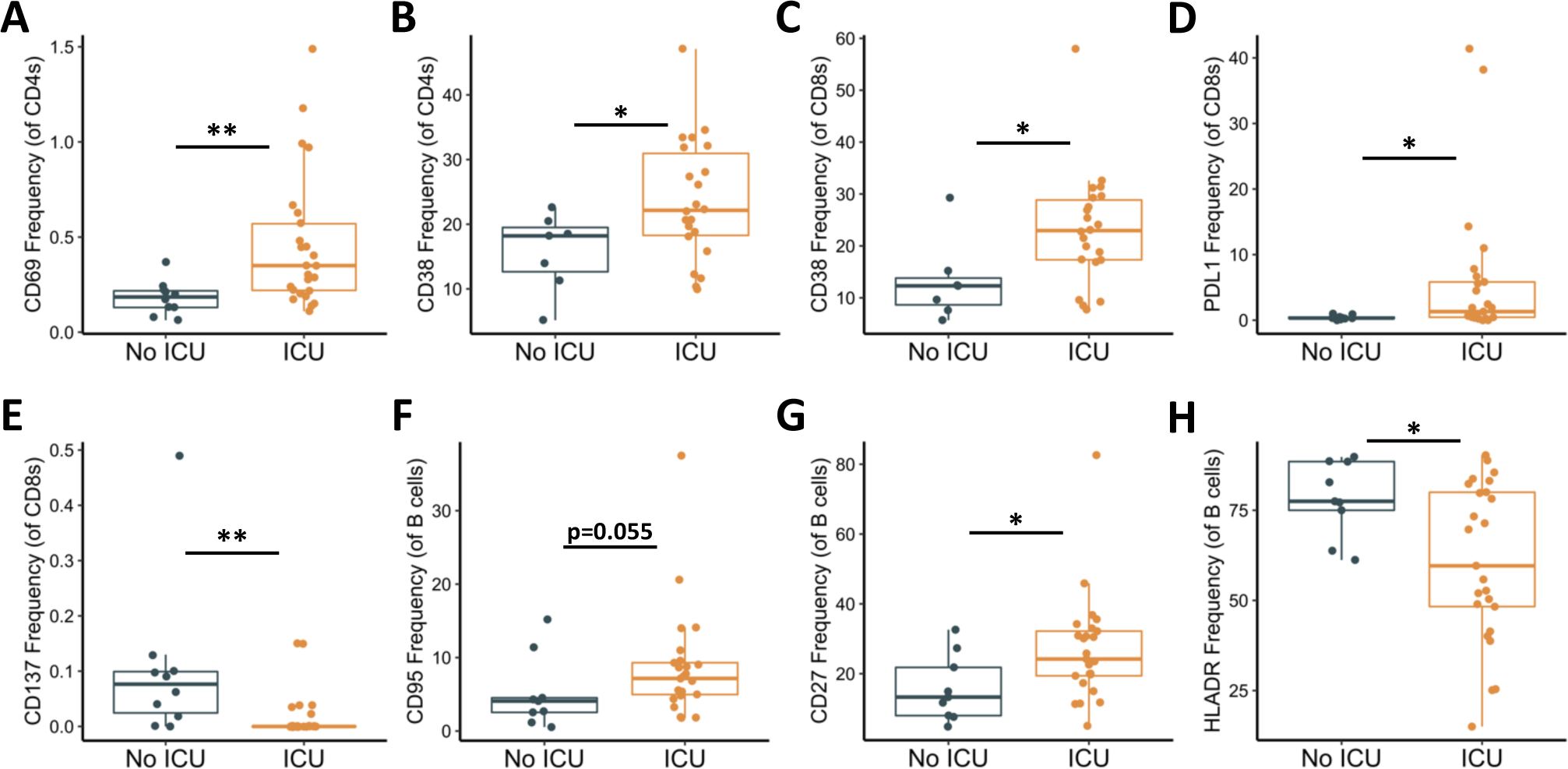
Activation and exhaustion markers in viremic hospitalized ICU patients. Frequencies of CD4+ T cells, CD8+ T cells, and B cells expressing given activation or exhaustion markers. **A-B)** Expression of CD69 and CD38 are elevated in CD4+ T cells of ICU patients. **C-D)** Expression of CD38 and PDL1 are elevated in CD8+ T cells, while **E)** expression of CD137 is decreased. **F-G)** CD95 and CD27 expression is increased in B cells, while **H)** HLA-DR expression is decreased. Boxplots indicate median, IQR, and 95% confidence interval. P-values determined by Wilcoxon rank sum tests and are indicated as follows: *p≤0.05, **p≤0.01, ***p≤0.001. n_No ICU_ = 10, n_ICU_= 26.

## DISCUSSION

Here we provide a comprehensive look at immune cell subsets during and after COVID-19 infection of hospitalized and non-hospitalized individuals. We found dysregulation of several immune cell types in hospitalized patients, whereas most had returned to baseline in non-hospitalized individuals. We also provide an in-depth characterization of the activation and exhaustion phenotype of CD4+ T cells, CD8+ T cells and B cells. While we observed activation marker upregulation in hospitalized patients, we also found several activation and exhaustion markers were expressed at higher frequencies in non-hospitalized convalescent samples. When investigating these markers, we observed several positive relationships over time, indicating that immune dysregulation in these individuals does not resolve quickly. We also found the dysregulation of T-cell activation and exhaustion markers in non-hospitalized individuals appears more pronounced in the elderly. To our knowledge, this is the first description of sustained immune dysregulation due to COVID-19 in a large group of non-hospitalized convalescent patients.

A recent study classified immune subsets in SARS-CoV-2 infection and reported increased frequencies of the classical CD14+ monocyte population in a small cohort of COVID-19 recovered patients (16), while a new report found increased frequencies of “Non-T/Non-B” cells in COVID-19 patients (26). Our results found decreased frequencies within monocytes, NK cells, and dendritic cells in hospitalized COVID-19 patients, that then returned to baseline in our non-hospitalized individuals. Taken together, these findings may suggest an influx of immature non-classical immune cells during infection and needs to be investigated further. Increased percentages of intermediate CD14+CD16+ monocytes in severe SARS-CoV-2 infections has been previously described (47). We found a trend (p=0.104) towards increased double positive monocytes in our hospitalized group, and also demonstrate that this monocyte population is elevated in non-hospitalized individuals. The intermediate monocyte population was recently shown to be functionally heterogenous (48), emphasizing the need for more investigation into this subset. Finally, we show that the frequency of dendritic cells is decreased in both hospitalized and non-hospitalized patients. Taken together, these data suggest a prolonged impact of SARS-CoV-2 on the innate immune system in both hospitalized and non-hospitalized individuals.

Our investigation into the CD4+ and CD8+ T-cell compartments clearly shows sustained activation (based on HLA-DR, CD69 and Ox40 expression) and exhaustion (based on PDL1 and TIGIT expression) in both T-cell subsets. As PD1 and Tim3 can be upregulated on both activated and exhausted T cells (34, 35), their role post COVID-19 infection remains unclear. Regulatory T cells have previously been shown to upregulate Ox40 and HLA-DR (49, 50). The sustained increase in Ox40 and HLA-DR in both hospitalized and non-hospitalized individuals could represent a regulatory T cell population, further emphasizing the need for further classification and functional assessment these subsets. The loss of CD27 and CD28 may suggest increased T-cell senescence, which is of particular relevance to understanding formation of memory responses.

CD4+ and CD8+ memory T cells are typically generated following the initial activation and expansion stage that occurs in acute infection (51). As previously demonstrated, establishment of airway memory CD4+ T cells mediated protective immunity against respiratory coronaviruses including SARS and MERS in a mouse model (52). Here we observe sustained expression of T-cell activation and exhaustion markers in non-hospitalized, convalescent SARS-CoV-2 individuals, as well as decreased frequencies of CD27 and CD28 expressing CD8+ T cells. These findings may represent an impaired ability to form memory T cells. This is supported by prior observations on IL-10 production in SARS and SARS-CoV-2 individuals where IL-10 production by regulatory CD4+ T cells was shown to be necessary for memory CD8+ T cell development in mice (53). In SARS patients, there was shown to been an increase in IL-10 production during the convalescent phase of infection (54); however, a recent report on SARS-CoV-2 infected individuals has found that serum levels of IL-10 were highest during acute infection and decreased in convalescence (15). Overall, these findings provide a possible etiology for how the sustained immune dysfunction observed during convalescence could impair the formation of long-term memory T cells, emphasizing the need to explore memory and regulatory T cell development and function in SARS-CoV-2 infected individuals.

There was substantial B cell activation demonstrated by increased frequencies of CD95+, CD69+, and PD1+ B cells in our hospitalized group. This may reflect both the presence of SARS-CoV-2 specific B cells responding to antigen and/or bystander B-cell activation. B-cell markers were found to generally return to levels similar to healthy controls, although the sustained presence of FcRL4+ and PD1+ B cells suggests persistence of some degree of B-cell dysregulation. How this dysregulation relates to SARS-CoV-2 antibody responses is unknown.

The finding of more pronounced T-cell activation/exhaustion in elderly non-hospitalized SARS-CoV-2 individuals has many potential implications. In acute disease, these findings suggest this group may be at heightened risk for inflammation-mediated pathology. This immune dysfunction may also lead to suboptimal SARS-CoV-2-specific memory responses and increased susceptibility of re-infection. Additional longitudinal studies are needed to better understand the impact of T-cell activation on long term immunity.

We extended our analysis to separate hospitalized patients that did or did not require intensive care. This revealed several differences in surface markers on T and B cell populations. Not surprisingly, activation markers were more frequently observed in severely infected individuals, in support of recent observations on the expression of HLA-DR and CD38 (26, 27). Our findings are the first report of upregulated PDL1 in severe COVID-19 patients. Interestingly, we observed that non-ICU patients had increased expression of CD137. Prior studies observed that CD137 can augment immune responses in acute viral infections, and that blocking CD137 with anti-CD137 antibodies led to increased LCMV infection in the mouse model (46). Our findings suggest CD137 expression may be associated with COVID-19 disease outcomes. Finally, we found that severely infected COVID-19 patients have increased B-cell expression of CD95 and CD27, and decreased expression of HLA-DR. These findings need to be investigated further to explore possible therapeutic interventions.

The prolonged activation we observed, particularly of T cells and monocytes, may be due to the persistence of antigen, either viral RNA and/or protein, despite resolution of symptoms. Support of this can be found from the studies that identified the persistence of viral RNA in SARS-CoV-2 patients during the convalescent period (55, 56). Antigen presenting cells (APCs) that present viral RNA can cross-prime and activate CD8+ T cells through a TLR3 mechanism (57), which could be similar to what is occurring in SARS-CoV-2 infected individuals. These data open the door to many interesting mechanistic studies to determine what factors are causing and prolonging immune cell activation.

Our current study has a few limitations. For one, our immunophenotyping panel prioritizes the identification of some subsets such as lymphocytes and monocytes over others, primarily dendritic cells. Although a marker for CD11c would have increased our ability to identify this population, we believe our gating strategy (CD3-CD19-CD14-CD56-CD16-HLADRhi) still identifies a relatively pure population of dendritic cells. Additionally, we acknowledge that there are substantial differences in age, race and sex between our hospitalized, non-hospitalized and healthy groups as summarized in **Table 1**. These differences reflect the nature of the COVID-19 pandemic, with numerous sources reporting increased hospitalizations and more severe clinical symptoms in elderly and African-American populations. We have controlled for this by performing general linear models comparing hospitalized, non-hospitalized, and healthy groups in pairwise models that have each been adjusted for participant age, race, and sex. Results are summarized in **Supplementary Tables 1-3**. This analysis shows that a majority of the significant relationships as determined by Wilcoxon analysis remain significant after adjusting for age, race and sex. In the remaining relationships that were not significant by hospitalization status, there were also no significant findings to support that these relationships were driven by age, race, or sex. One exception to this was the finding that the decrease in CD16+ monocytes in our hospitalized group appears to be driven by sex (**Supplemental Table 1**). In addition, our hospitalized group had an increased frequency of existing comorbidities (shown in **Table 1**). This was expected since many of these risk factors have been found to correlate with worsening clinical outcomes in COVID-19 infection. Our study provides important insights into acute and subacute COVID-19 immune responses but leaves questions unanswered in relation to comorbidities and their impact on immunophenotyping. Future studies using larger cohorts should directly examine the potential impact of these comorbidities on immune cell dysregulation in COVID-19 infection.

In conclusion, this study provides broad insight into the regulation of several immune cell subsets and identifies immune cell dysregulation in both hospitalized and non-hospitalized SARS-CoV-2 infected individuals. How long T-cell and B-cell dysregulation persists after COVID-19 infection and whether this may alter immune responsiveness to subsequent infectious insults is not yet clear. Similarly, sustained B-cell and T-cell activation may have consequences for the development or exacerbation of other inflammatory diseases. These results highlight the need for additional studies in COVID-19 patients to further define the immune landscape in these individuals.

## METHODS

### Sample Collection

Peripheral blood samples were collected from hospitalized (n=46) and non-hospitalized (n=39) patients at the University of Alabama at Birmingham. Samples were also collected from 25 non-hospitalized samples at a second time point. Convalescent status was defined based on patients being asymptomatic for at least 3 consecutive days and being at least 7 days past initial diagnosis. Patient clinical data for hospitalized patients was collected from the electronic medical record; self-reported clinical data was collected for convalescent samples by questionnaire at the time of sample collection and uploaded to REDCap (58). All data and samples were collected in accordance with the University of Alabama at Birmingham’s IRB. All patients had a confirmed positive test for SARS-CoV-2 unless otherwise stated. Summary of patient demographic and clinical status data is shown in **Table 1**. Peripheral blood mononuclear cells (PBMCs) from patient blood samples were harvested using density gradient centrifugation. Additionally, our cohort of healthy (CoV-) individuals were comprised of 20 total samples. Of these, frozen PBMCs (n=17) from before 2019 were used primarily to ensure the absence of asymptomatic SARS-CoV-2 infections; additionally, fresh samples were collected from three asymptomatic seronegative individuals, and analysis from each of these samples showed no significant differences between fresh and frozen samples within that individual.

### Flow Cytometric Analyses

Isolated PBMCs were stained using one of four different phenotyping panels (**Supplemental Table 4**). Immune cell subsets were identified with the following immunophenotyping panel: CD16-FITC, CD14-A700, CD45-Pecy7, CD19-Percpcy5.5, CD27-PEAlexa610, CD56-BV421, CD3-A780, CD8-V500, CD4-BV785, and HLADR-PE. T cells were further characterized using two additional panels: (1) CD8-V500, CD3-BV711, CD4-BV786, CD38-BUV737, CD28-APC; and (2) TIGIT-PerCpCy5.5, PDL1-PE, CD4-PEAlexa610, Ox40-Pecy7, Tim3-BV421, CD8-V500, CD137-BV650, PD1-BV785, CD14-BUV563, CD19-BUV563, CD154-APC, CD3-A780. B cell phenotype was assessed using the following panel: (a) CD4-BB790, CD19-AF700, FcRL4-BV480, CD27-BV650, CD69-BUV395, CD8-BUV496, PD1-BUV563, HLA-DR-BUV661, CD95-BUV737. All panels utilized Live/Dead Blue Stain (Life Technologies) to identify dead cells. After staining, samples were fixed using a 1% formalin solution. Events were collected using a FACS Symphony A3 (BD Biosciences) flow cytometer and analyzed using FlowJo (v10, Treestar) software. Representative gating strategies can be seen in **Supplemental Figures 5-8**.

### Statistics

All statistical analysis and figure generation was performed using R. Significance between groups was performed using Wilcoxon rank sum test (unpaired) or Wilcoxon signed rank test (paired). Significance between two continuous variables was calculated by using a Spearman’s correlation test or a mixed effect model in order to account for convalescent individuals with multiple time points. Additionally, linear modeling was used to verify significant relationships between different hospitalization groups were not driven by differences in age, race, or sex.

## Data Availability

Data were generated at University of Alabama at BIrmingham. Derived data supporting the findings of this study are available from the corresponding authors (NE/PAG) on request.

## AUTHOR CONTRIBUTIONS

Both JKF and MDP performed staining assays for all fresh samples. JKF performed initial flow cytometric immunophenotyping data collection and analysis; JKF/MDP/KEL performed initial flow cytometric T-cell data collection and analysis; S. Sarkar (SS^1^) performed initial flow cytometric B-cell data collection and analysis. SB and JKF performed finalized data analyses and statistical tests, accessed most clinical information, and generated all figures. JKF primarily wrote the paper, receiving assistance from KQ/MDP/KEL/SS^1^ for the introduction, assistance from SB for the results, and assistance from SB/MDP/KEL/JK for the discussion; all authors provided edits/feedback. S. Sterrett (SS^2^) helped provide initial clinical data. JKF, SB, KQ, SS^2^, EC, and JK all assisted with PBMC processing. AB and S. Sabbaj (SS^3^) provided critical scientific expertise and comments. OK, JK, PAG, and NE all provided scientific knowledge, as well as supplies including the required flow cytometry antibodies. PAG and NE recruited patients and obtained all clinical samples.

## ACKNOWLEDGEMENTS

We would first like to thank all patients for giving consent for sample collection for this study. Additionally, we want to thank the clinical collection and UAB CFAR biorepository teams for their assistance in sample collection, as well as the hospital staff for their dedication in fighting coronavirus. We would like to acknowledge Lynn Pritchard, Tim Fram, and David Moylan who all processed several PBMC samples used in this project, as well as the UAB CFAR Basic Research Core (P30 AI027767-31) for use of flow cytometer and BSL2+ facility. Finally, we would also like to acknowledge Anisha Jackson and Ainsley Greenstein for help in obtaining clinical data.

## REFERENCES

1. Zhu N, Zhang D, Wang W, Li X, Yang B, Song J, et al. A Novel Coronavirus from Patients with Pneumonia in China, 2019. N Engl J Med. 2020;382(8):727–33.

2. Li Q, Guan X, Wu P, Wang X, Zhou L, Tong Y, et al. Early Transmission Dynamics in Wuhan, China, of Novel Coronavirus-Infected Pneumonia. N Engl J Med. 2020;382(13):1199–207.

3. Wang W, Xu Y, Gao R, Lu R, Han K, Wu G, et al. Detection of SARS-CoV-2 in Different Types of Clinical Specimens. JAMA. 2020.

4. Huang C, Wang Y, Li X, Ren L, Zhao J, Hu Y, et al. Clinical features of patients infected with 2019 novel coronavirus in Wuhan, China. Lancet. 2020;395(10223):497–506.

5. Richardson S, Hirsch I, Narasimhan M, Crawford JM, McGinn T, and Davidson KW, et. al. Presenting Characteristics, Comorbidities, and Outcomes Among 5700 Patients Hospitalized With COVID-19 in the New York City Area. JAMA. 2020.

6. Chen N, Zhou M, Dong X, Qu J, Gong F, Han Y, et al. Epidemiological and clinical characteristics of 99 cases of 2019 novel coronavirus pneumonia in Wuhan, China: a descriptive study. Lancet. 2020.

7. Johns Hopkins Coronavirus Resource Center https://coronavirus.jhu.edu/. Updated May 13, 2020 Accessed May 13, 2020.

8. Wang F, Nie J, Wang H, Zhao Q, Xiong Y, Deng L, et al. Characteristics of peripheral lymphocyte subset alteration in COVID-19 pneumonia. J Infect Dis. 2020.

9. Guan WJ, Ni ZY, Hu Y, Liang WH, Ou CQ, He JX, et al. Clinical Characteristics of Coronavirus Disease 2019 in China. N Engl J Med. 2020;382(18):1708–20.

10. Wang D, Hu B, Hu C, Zhu F, Liu X, Zhang J, et al. Clinical Characteristics of 138 Hospitalized Patients With 2019 Novel Coronavirus–Infected Pneumonia in Wuhan, China. JAMA. 2020.

11. Chen G, Wu D, Guo W, Cao Y, Huang D, Wang H, et al. Clinical and immunological features of severe and moderate coronavirus disease 2019. Journal of Clinical Investigation. 2020.

12. Zheng HY, Zhang M, Yang CX, Zhang N, Wang XC, Yang XP, et al. Elevated exhaustion levels and reduced functional diversity of T cells in peripheral blood may predict severe progression in COVID-19 patients. Cell Mol Immunol. 2020.

13. Zheng M, Gao Y, Wang G, Song G, Liu S, Sun D, et al. Functional exhaustion of antiviral lymphocytes in COVID-19 patients. Cell Mol Immunol. 2020.

14. Ong EZ, Chan FZC, Leong WY, Lee NMY, Kalimuddin S, Mohideen SMH, et al. A Dynamic Immune Response Shapes COVID-19 Progression. Cell Host & Microbe. 2020.

15. Diao B, Wang C, Tan Y, Chen X, Liu Y, Ning L, et al. Reduction and Functional Exhaustion of T Cells in Patients with Coronavirus Disease 2019 (COVID-19). Frontiers of Immunology. 2020;11:827.

16. Wen W, Su W, Tang H, Le W, Zhang X, Zheng Y, et al. Immune cell profiling of COVID-19 patients in the recovery stage by single-cell sequencing. Cell Discov. 2020;6:31.

17. Grifoni A, Weiskopf D, Ramirez SI, Mateus J, Dan JM, Moderbacher CR, et al. Targets of T cell responses to SARS-CoV-2 coronavirus in humans with COVID-19 disease and unexposed individuals. Cell. 2020.

18. Ni L, Ye F, Cheng ML, Feng Y, Deng YQ, Zhao H, et al. Detection of SARS-CoV-2-Specific Humoral and Cellular Immunity in COVID-19 Convalescent Individuals. Immunity. 2020.

19. Assiri A, Al-Tawfi JA, Al-Rabeeah AA, Al-Rabiah FA, Al-Hajjar S, Al-Barrak A, et al. Epidemiological, demographic, and clinical characteristics of 47 cases of Middle East respiratory syndrome coronavirus disease from Saudi Arabia: a descriptive study. The Lancet: Infectious Diseases. 2013.

20. Yin Y, and Wunderink RG. MERS, SARS and other coronaviruses as causes of pneumonia. Respirology. 2018.

21. Chen H, Hou J, Jiang X, Ma S, Meng M, Wang B, et al. Response of memory CD8+ T cells to severe acute respiratory syndrome (SARS) coronavirus in recovered SARS patients and healthy individuals. J Immunol. 2005;175(1):591–8.

22. Chen J, and Subbarao K. The Immunobiology of SARS‪. Annu Rev Immunol. 2007;25:443–72.

23. Zhao J, Zhao J, and Perlman S. T cell responses are required for protection from clinical disease and for virus clearance in severe acute respiratory syndrome coronavirus-infected mice. J Virol. 2010;84(18):9318–25.

24. Wang F, Hou H, Luo Y, Tang G, Wu S, Huang M, et al. The laboratory tests and host immunity of COVID-19 patients with different severity of illness. JCI Insight. 2020.

25. Thevarajan I, Nguyen THO, Koutsakos M, Druce J, Caly L, van de Sandt CE, et al. Breadth of concomitant immune responses prior to patient recovery: a case report of non-severe COVID-19. Nature Medicine. 2020.

26. Mathew D, Giles JR, Baxter AE, Derek AO, Greenplate AR, Wu JE, et al. Deep immune profiling of COVID-19 patients reveals distinct immunotypes with therapeutic implications. Science. 2020.

27. Kuri-Cervantes L, Pampena MB, Meng W, Rosenfeld AM, Ittner CAG, Weisman AR, et al. Comprehensive mapping of immune perturbations associated with severe COVID-19. Science Immunology. 2020.

28. Wang Z, Zhu L, Nguyen T, Wan Y, Sant S, Quiñones-Parra Sm, et al. Clonally diverse CD38+HLA-DR+CD8+ T cells persist during fatal H7N9 disease. Nature Communications. 2018;9(1):824.

29. Isa A, Kasprowicz V, Norbeck O, Loughry A, Jeffery K, Broliden K, et al. Prolonged Activation of Virus-Specific CD8T Cells after Acute B19 Infection. PloS Medicine. 2005.

30. Wong SS, Oshansky CM, Guo XZJ, Ralston J, Wood T, Seeds R, et al. Severe Influenza Is Characterized by Prolonged Immune Activation: Results From the SHIVERS Cohort Study. Journal of Infectious Diseases. 2018.

31. Choe PG, Perera Rapm, Park WB, Song KH, Bang JH, Kim ES, et al. MERS-CoV Antibody Responses 1 Year after Symptom Onset, South Korea, 2015. Emerging Infectious Disease. 2017.

32. Chen T, Wu D, Chen H, Yan W, Yang D, Chen G, et al. Clinical characteristics of 113 deceased patients with coronavirus disease 2019: retrospective study. BMJ. 2020;368:m1091.

33. Scharenberg M, Vangeti S, Kekäläinen E, Bergman P, Al-Ameri M, Johansson N, et al. Influenza A Virus Infection Induces Hyperresponsiveness in Human Lung Tissue-Resident and Peripheral Blood NK Cells. Frontiers of Immunology. 2019.

34. Gorman JV, and Colgan JD. Regulation of T cell responses by the receptor molecule Tim-3. Immunologic Research. 2014.

35. Jubel J, Barbarti Z, Burger C, Wirtz D, and Schildberg F. The Role of PD-1 in Acute and Chronic Infection. Frontiers of Immunology. 2020.

36. Effros RB. Loss of CD28 expression on T lymphocytes: a marker of replicative senescence. Dev Comp Immunol. 1997.

37. Jourdan M, Robert N, Cren M, Thibaut C, Duperray C, Kassambara A, et al.Characterization of human FCRL4-positive B cells. PloS One. 2017.

38. Weiss GE, Crompton PD, Li S, Walsh LA, Moir S, Traore B, et al. Atypical Memory B Cells Are Greatly Expanded in Individuals Living in a Malaria-Endemic Area. Journal of Immunology. 2009.

39. Poonia B., Ayithan N., Nandi M., Masur H., and S. K. HBV induces inhibitory FcRL receptor on B cells and dysregulates B cell-T follicular helper cell axis.. Scientific Reports. 2018.

40. Moir S, Ho J, Malaspina A, Wang W, DiPoto AC, O’Shea MA, et al. Evidence for HIV-associated B cell exhaustion in a dysfunctional memory B cell compartment in HIV-infected viremic individuals Journal of Experimental Medicine. 2008.

41. Amara K, Clay E, Yeo L, Ramskold D, Spengler J, Sippl N, et al. B cells expressing the IgA receptor FcRL4 participate in theautoimmune response in patients with rheumatoid arthritis. Journal of Autoimmunity. 2017.

42. Ehrhardt GRA, Hsu JT, Gartland L, Leu CM, Zhang S, Davis RS, et al. Expression of the immunoregulatory molecule FcRH4 defines a distinctive tissue-based population of memory B cells. Journal of Experimental Medicine. 2005.

43. Wang D., Hu B., Hu C., Zhu F, Liu X, Zhang J, et al. Clinical Characteristics of 138 Hospitalized Patients With 2019 Novel Coronavirus–Infected Pneumonia in Wuhan, China. JAMA. 2020.

44. Sterrett S, Peng BJ, Burton RL, LaFon DC, Westfall AO, Singh S, et al. Peripheral CD4 T follicular cells induced by a conjugated pneumococcal vaccine correlate with enhanced opsonophagocytic antibody responses in younger individuals. Vaccine. 2020.

45. Chong Y, Ikematsu H, Yamaji K, Nishimura M, Nabeshima S, Kashiwagi S, et al. CD27+ (memory) B cell decrease and apoptosis-resistant CD27-(naive) B cell increase in aged humans: implications for age-related peripheral B cell developmental disturbances. International Immunology. 2005.

46. Zhang B, Mais CH, Foell J, Whitmire J, Niu L, Song J, et al. Immune suppression or enhancement by CD137 T cell costimulation during acute viral infection is time dependent. JCI. 2007.

47. Zhou Y, Fu B, Zheng X, Wang D, Zhao C, qi Y, et al. Pathogenic T cells and inflammatory monocytes incite inflammatory storm in severe COVID-19 patients. Natl Sci Rev. 2020.

48. Villani A, Satija R, Reynolds G, Sarkizova S, Shekhar K, Fletcher J, et al. Single-cell RNA-seq reveals new types of human blood dendritic cells, monocytes, and progenitors. Science. 2017.

49. Reiss S., Baxter A.E., Cirelli K.M., Dan J.M., Morou A., Daigneault A., et al. Comparative analysis of activation induced marker (AIM) assays for sensitive identification of antigen-specific CD4 T cells.. PLos ONE. 2017.

50. Machicote A, Belén S, Baz P, La B, and Fainboim L. Human CD8+ HLA-DR+ Regulatory T Cells, Similarly to Classical CD4+ Foxp3+ Cells, Suppress Immune Responses via PD-1/PD-L1 Axis. Frontiers of Immunology. 2018.

51. Wherry EJ, and Ahmed R. Memory CD8 T-Cell Differentiation during Viral Infection.Journal of Virology. 2004.

52. Zhao J, Zhao J, Mangalam AK, Channappanavar R, Fett C, Meyerholz DK, et al. Airway Memory CD4(+) T Cells Mediate Protective Immunity against Emerging Respiratory Coronaviruses. Immunity. 2016;44(6):1379–91.

53. Laidlaw BJ, Cui W, Amezquita RA, Gray SM, Guan T, Lu Y, et al. Production of IL-10 by CD4(+) regulatory T cells during the resolution of infection promotes the maturation of memory CD8(+) T cells. Nature Immunology. 2015.

54. Zhang Y, Li J, Zhan Y, Wu L, Yu X, Zhang W, et al. Analysis of serum cytokines in patients with severe acute respiratory syndrome. Infection and Immunity. 2004.

55. Chen D, Xu W, Lei Z, Huang Z, and Liu J. Recurrence of positive SARS-CoV-2 RNA in COVID-19: A case report. International Journal of Infectious Diseases. 2020.

56. Wölfel R, Corman VM, Guggemos W, Seilmaier M, Zange S, Müller MA, et al. Virological assessment of hospitalized patients with COVID-2019. Nature. 2020.

57. Schulz O, Diebold S, Chen M, Näslund TI, Nolte MA, Alexopoulou L, et al. Toll-like receptor 3 promotes cross-priming to virus-infected cells. Nature. 2005(433):887–92.

58. PA Harris, R Taylor, BL Minor, V Elliott, M Fernandez, L O’Neal, et al. The REDCap consortium: Building an international community of software partners. J Biomed Inform. 2019.

